# Packed Red Blood Cell and Whole Blood Perfusates during an Ex-vivo Normothermic Perfusion for Assessment of High-Risk Donor Kidneys

**DOI:** 10.1101/2024.10.18.24315776

**Authors:** Armin Ahmadi, Heiko Yang, Kuang-Yu Jen, Sili Fan, Ivonne Palma, Junichiro Sageshima, Naeem Goussous, Baback Roshanravan, Richard V. Perez

## Abstract

**Background:** Ex-vivo normothermic perfusion (EVNP) with a blood based perfusate has the potential to both assess viability of and repair high-risk organs prior to transplantation. However, the optimal perfusate is yet to be established.

**Methods:** Eight paired high-risk human kidneys were placed on three hours of pressure dependent EVNP at 37°C. Kidneys were perfused with either a leukocyte-depleted packed red blood cell (PRBC) with crystalloid perfusate or a whole blood (WB) perfusate. Continuous hemodynamic and functional parameters were assessed. Core needle tissue biopsies were taken for untargeted metabolomics and lipidomics profiling. A *t test* was used to assess group differences in functional and hemodynamics outcomes.

**Results:** After a mean cold ischemia time (CIT) of 54 hours, all kidneys showed high renal blood flow (RBF) through perfusion. Renal resistance (RR) increased for both groups during the first hour and then decreased to similar terminal values. The kidneys perfused with PRBC had 55 ml/min greater RBF (95% CI of 21 to 89; P=0.004) and higher total urine output (UO) (145 vs 25 ml, P= 0.002) compared to the WB group. Urinary acute kidney biomarkers of NGAL and KIM-1 were also significantly lower (mean differences of 281 and 2.1 ng/ml respectively; P<0.01) in the PRBC perfused kidneys. Compared to PRBC, within group tissue metabolic profiling revealed a similar (23% vs 18%) but a more pronounced alteration predominantly involving (branched chain) amino acid and mitochondrial energy metabolism in the WB group. Similarly, lipid profile temporal changes showed WB group were highlighted by elevation of plasma membrane and structure lipids including glycerolipids, sphingolipids, and steroids. The PRBC group had minimal temporal tissue lipid profile changes.

**Conclusion:** Compared to WB, PRBC perfusion is superior in mitigating post-ischemia damage and facilitating function and metabolic recovery of high-risk kidneys subjected to long CITs during a three-hour ENVP.

**Key points:**

- Kidneys perfused with packed red blood cells (PRBC) exhibited more favorable hemodynamics and functional outcomes during ex vivo normothermic perfusion (ENVP).
- Tissue metabolic profiling showed an overlapping but a more drastic metabolic aberration in the whole blood (WB) group involving amino acid, carbohydrate, and mitochondrial energy metabolism over time.
- Compared to the PRBC groups, WB perfused kidneys were associated with accumulation of tissue membrane/structure components including TGs, ceramides, and cholesteryl esters post perfusion.

## Introduction

The most significant obstacle in kidney transplantation is the shortage of available donor organs^1^. To address the shortage, the donor pool has been expanded to include higher risk organs such as those with acute kidney injury, donation after circulatory death, and older donors with defined comorbidities previously termed extended criteria donors. While improved survival has been demonstrated after transplantation with these higher risk organs^2^, many initially recovered for transplantation are discarded due to the lack of optimal perfusion to preserve and facilitate functional recovery before transplantation ^3,4^.

Ex vivo normothermic perfusion (EVNP) has been reported as a means of preserving and assessing the viability of high-risk kidneys by restoring metabolic physiological conditions prior to transplantation^5,6^. With re-establishment of normal metabolism using a blood based perfusate, EVNP can replenish mitochondrial energy stores with increased adenosine triphosphate levels^7^ and improve renal allograft function after ischemia induced injury^6,8,9^. The majority of current kidney perfusion protocols involve continuous perfusion with red blood cells or an oxygen carrier alternative and glucose and amino acids as energy sources. However, the optimal perfusate for use during EVNP to efficiently improve viability, function, and metabolic health is yet to be determined.

A packed red blood cell (PRBC)-based has been used in most studies due to evidence suggesting that leukocytes and platelets contribute to ischemia reperfusion injury^10-13^. Indeed, mechanistic studies have linked the presence of leukocytes and platelets with an amplified ischemia reperfusion injury response including heightened levels of ROS generation, increased neutrophil infiltration, and tubular and endothelial damage^14^. On the other hand, whole blood (WB) has been shown to be superior to a PRBC perfusate in an experimental model of heart transplantation presumably due to its more accurate reproduction of normal physiologic conditions^15^. Here, we performed a three-hour ENVP during which we assessed various kidney hemodynamics and functional parameters, and we also performed untargeted metabolomics and lipidomics using kidney cortical tissues from 8 paired kidneys. The purpose of this study was twofold: first, to compare the impact of WB and PRBC perfusate solutions on restoring function of high-risk kidneys and second, to characterize their respective metabolic implications during a three-hour ENVP period.

## Methods

### Deceased donor kidneys

The study was reviewed by and considered exempt from the Institutional Review Board at our institution and approved by the Human Anatomical Specimen and Tissue Oversight Committee. In all cases, consent for research was obtained from the donor family and organ procurement organization. Eight paired human kidneys from four donors deemed unsuitable for transplantation after being offered and declined were included in this study and transported to our research lab at the University of California Davis Health System.

### EVNP system

All kidneys were placed on an EVNP system using a pediatric cardiopulmonary bypass system as previously described^16^. Once connected to the circuit, the kidneys were continuously perfused between systolic pressure of 70 to 80 mmHg for three hours at a temperature of 37° and flow was adjusted to keep pressure between set points.

Paired kidneys from each donor were either perfused with a PRBC or WB based perfusate (n= 8). The PRBC perfusate consisted of leukocyte and platelet depleted, washed red cell unit blood type O positive, which were acquired from our internal hospital blood bank and diluted in a 1:1 ratio with Plasmalyte-A (Baxter Medical, Deerfield, IL). The WB perfusate was type specific blood in citrate phosphate dextrose adenine solution, collected 24 hours prior from the local blood center laboratory from repeated male donors with no history of infectious diseases stored at room temperature (Bloodsource, Mather, CA).

The perfusates were supplemented with heparin (2000 international units, NOVAPLUS), exogenous anhydrous creatinine (0.06 g, MP Biomedicals, Burlingame, CA), 20 ml of parenteral nutrition (Baxter CLINIMIX E 2.75/10) which was infused with 100 units of regular insulin (Humulin R), 5 ml of multivitamins (Baxter Medical), and 26 ml of 8.4% Bicarbonate (Millipore Sigma, St. Louis, MO). The circuit was primed by re-circulation of the prepared PRBC or WB perfusate oxygenated with 95% O_2_ /5% CO_2_ gas mixture for one to two minutes. Urine loss was replaced at a 1:1 ratio with Plasmalyte-A for both perfusate solutions.

### Kidney assessment

Hemodynamic parameters (pressure and flow) were recorded every 30 minutes and renal resistance (RR) was calculated as pressure (mmHg)/ flow (ml/min). Urine was collected every 30 minutes and stored in for further analysis. Urine samples were later analyzed for sodium using a Critical Care Xpress Machine (Nova Biomedical, Waltham, MA) and for creatinine using a creatinine parameter assay kit (R&D systems, Minneapolis, MN) for calculation of creatinine clearance and fractional excretion of sodium (FENa) as reported previously^16^.

Perfusate samples were collected every 30 minutes from a port in the arterial line and directly from the venous return. The samples were analyzed by a Critical Care Xpress machine for pH, pCO_2_, and electrolytes, and handheld point of care devices (Stat sensors, Nova Biomedical) for creatinine, lactate, and glucose levels. A wedge biopsy of the renal cortex was collected at the end of the perfusion for histology.

### Urinary acute kidney injury biomarkers

Urinary neutrophil gelatinase-associated lipocalin (NGAL) was measured with a point of care fluorescence immunoassay system (Bio Site Triage MeterPro Alere, Waltham, MA). Urinary kidney injury molecule-1 (KIM-1) was measured with a human urinary KIM-1 quantikine enzyme-linked immunosorbent assay kit per manufacturer’s instructions (R&D Systems).

### Metabolomics and Lipid Profile

Renal cortical core tissue needle biopsies (16g Tru Core II Biopsy Instrument, Argon Medical Devices, Athens, TX) were taken prior to and after EVNP and snap frozen in liquid nitrogen. After study completion, samples (n=16) were submitted to the West Coast Metabolomics Center at the University of California Davis Genome Center. Samples were analyzed with validated chromatography-mass spectrometry methods with 15-25 internal standards, for a non-targeted primary metabolites analysis. Lipidomics profiling was performed using liquid chromatography coupled to quadrupole time-of-flight mass spectrometer-charged surface hybrid (LC QTOF CHS). A total of 185 metabolites and 220 lipid species were identified across all 8 kidneys.

### Statistical Analysis

We used both *t* test and the area under the curve (AUC) measurements (using the trapezoid rule) to assess and validate group differences in RBF, RR, UO, perfusate creatinine, perfusate lactic acid, urinary NGAL, urinary KIM-1, and calculated FENa during ENVP. Additionally, we fit a linear mixed-effects model to compare changes over time during the three-hour ENVP. These models included a random effect for intercept and slope to accommodate for differences at baseline levels, dependency within matched pairs, and the repeated measures over time. We used analysis of variance adjusted for multiple hypothesis testing (Bonferroni) to determine significant differences in hemodynamics and metabolites over time. The means and standard error of mean are provided. We used a paired *t* test to assess within group tissue temporal metabolic and lipid profile changes. Similarly, baseline metabolic and lipid profile differences were assessed using a paired *t* test. To assess temporal tissue metabolic and lipid profile differences between the two groups a two-way ANOVA using the interaction of time (pre vs post) and group (PRBC vs WB) was used. MetaboAnalyst 6.0 was used for enrichment analysis. All tests were two-sides and P< 0.05 were concluded statistically significant. Analyses were conducted in R version 4.2.2.

## Results

### Donor demographics and characteristics of kidneys

The mean donor age was 61 years (range 54-67 years), and mean donor weight was 92 kg (range 66-137 kg) (**Table 1**). There was one DCD donor and three ECD donors with a mean kidney donor profile index of 87% (range 77-99%). Kidneys were discarded due to a combination of donor history, cold ischemia time (CIT), and biopsy results. Four of the paired kidneys were placed on HMP prior to discard for a mean time of 4 hours (**Table 1**).

**Table 1.** Donor demographics and characteristics of kidneys placed on ex-vivo normothermic perfusion. AS, arteriolosclerosis; CIT, cold ischemia time; CVA, Cerebrovascular Accident; DBD, donation after brain death; DCD, donation after cardiac death; DM, diabetes mellitus; ECD, extended criteria donor; F, female; GS, glomerulosclerosis; HTN, hypertension; IF, interstitial fibrosis; KDPI, kidney donor profile index; M, male; PRBC, packed red blood cell; SCD, standard criteria donor; TA, tubular atrophy; WB, whole blood.

| Kidney ID | 1 |  | 2 |  | 3 |  | 4 |  |
| --- | --- | --- | --- | --- | --- | --- | --- | --- |
| Age (yrs) | 67 |  | 58 |  | 54 |  | 64 |  |
| M/F | F |  | M |  | M |  | F |  |
| HTN | Yes |  | No |  | No |  | Yes |  |
| DM | Yes |  | No |  | No |  | Yes |  |
| Weight (kg) | 74 |  | 137 |  | 66 |  | 91 |  |
| Cause of Death | Anoxia |  | CVA |  | Anoxia |  | CVA |  |
| Terminal Creatinine (mg/dL) | 1.70 |  | 3.40 |  | 1.66 |  | 0.55 |  |
| KDPI (%) | 99 |  | 79 |  | 75 |  | 94 |  |
| ECD or SCD | ECD |  | ECD |  | SCD |  | ECD |  |
| DBD or DCD | DBD |  | DBD |  | DCD |  | DBD |  |
| Total CIT (hours) | 40 | 41 | 54 | 55 | 56 | 56 | 63 | 64 |
| Static Cold Storage (hrs) | 40 | 41 | 54 | 55 | 50 | 49 | 62 | 63 |
| Hypothermic Machine Perfusion (hrs) | 0 | 0 | 0 | 0 | 6 | 7 | 1 | 1 |
| Anatomical Position | Left | Right | Right | Left | Left | Right | Left | Right |
| Biopsy Results | 17% GS<br>Mild IF<br>Mild TA<br>Mild AS | 19% GS<br>Mild IF<br>Mild TA<br>Mild AS | 19% GS<br>Mild to<br>Mod IF<br>Mild TA<br>Mod to<br>Severe AS | 20% GS<br>Mild to<br>Mod IF<br>Mild to<br>Mod TA<br>Mod to<br>Severe AS | 12% GS<br>Mild IF<br>Mild TA<br>Mod AS<br>Benign<br>Cortical<br>Cyst | 14% GS<br>Mild IF<br>Mild TA<br>Mod AS | 0% GS<br>Mild IF<br>Mild TA<br>Severe AS | 0% GS<br>Mild IF<br>Mild TA<br>Mod to<br>Severe<br>AS |
| EVNP Perfusate | WB | PRBC | WB | PRBC | WB | PRBC | WB | PRBC |

### Hemodynamics and functional assessment

All kidneys exhibited a healthy pink color after the initiation of EVNP. The PRBC perfused kidney group initially had a higher mean RBF of 217 (77) ml/min compared to an initial flow of 150 (70) ml/min in the WB perfused kidney group and maintained a higher RBF throughout the perfusion period (**Figure 1A, Table 2**). RBF decreased in both groups to a nadir at one hour after which flow increased gradually thereafter until the end of perfusion period. Overall, RR was lower among the PRBC group, initially increasing over the first hour, followed by a gradual decrease for both groups (**Figure 1B, Table 2)**. Total urine output volume after EVNP was significantly higher in the PRBC group compared to the WB group (**Figure 1C, Table 2**).

**Figure 1.**
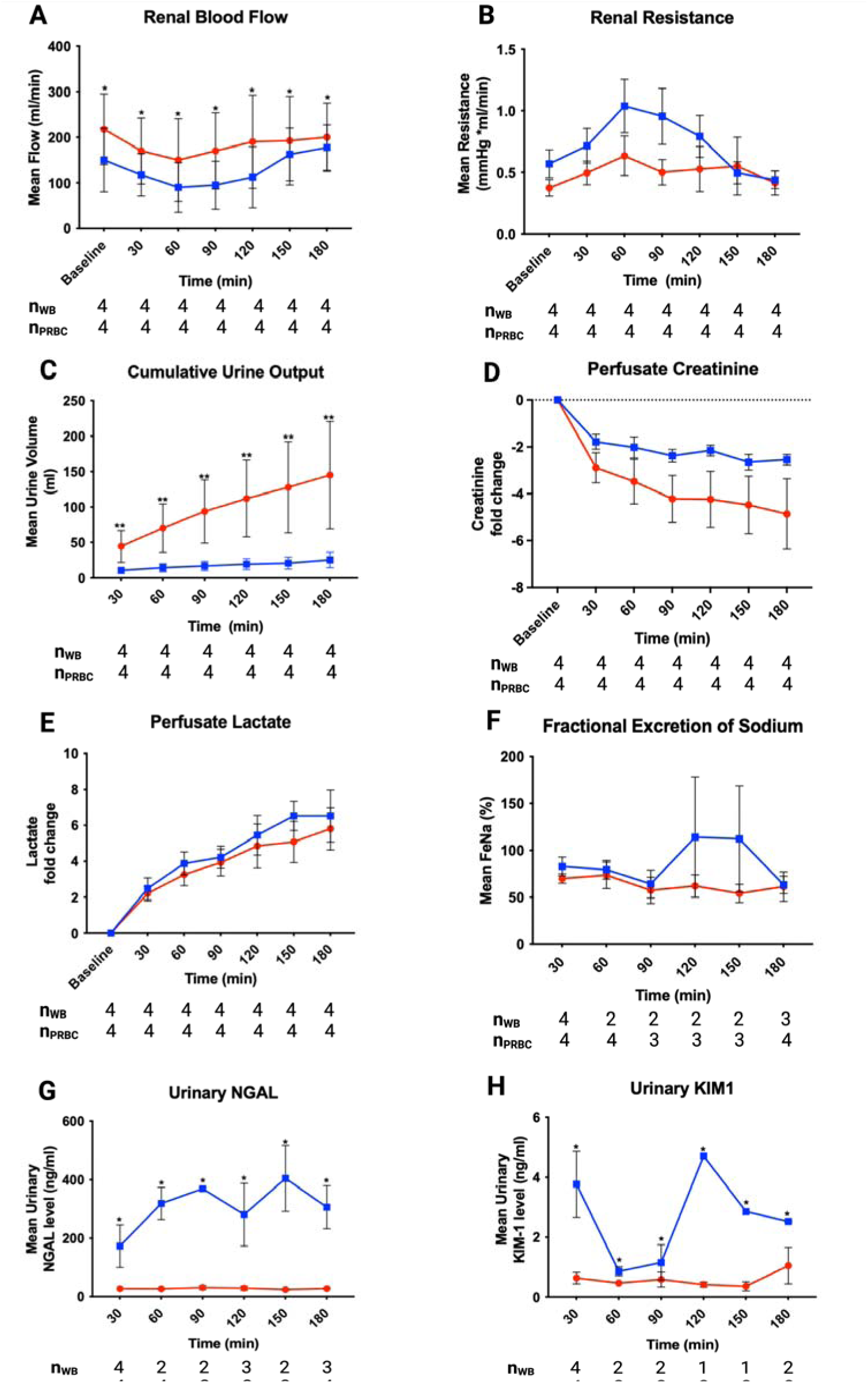
Hemodynamic and functional parameters as well as urinary biomarkers are compared over time within and between perfusate groups. Graphs are plotted as mean and error bars represent standard error of mean. The blue lines represent the WB group, and the red lines represent the PRBC group. The number of available data for each group at a given time point is shown at the bottom of each panel. ∗ P < 0.05; ∗∗ P < 0.001.

**Table 2.**
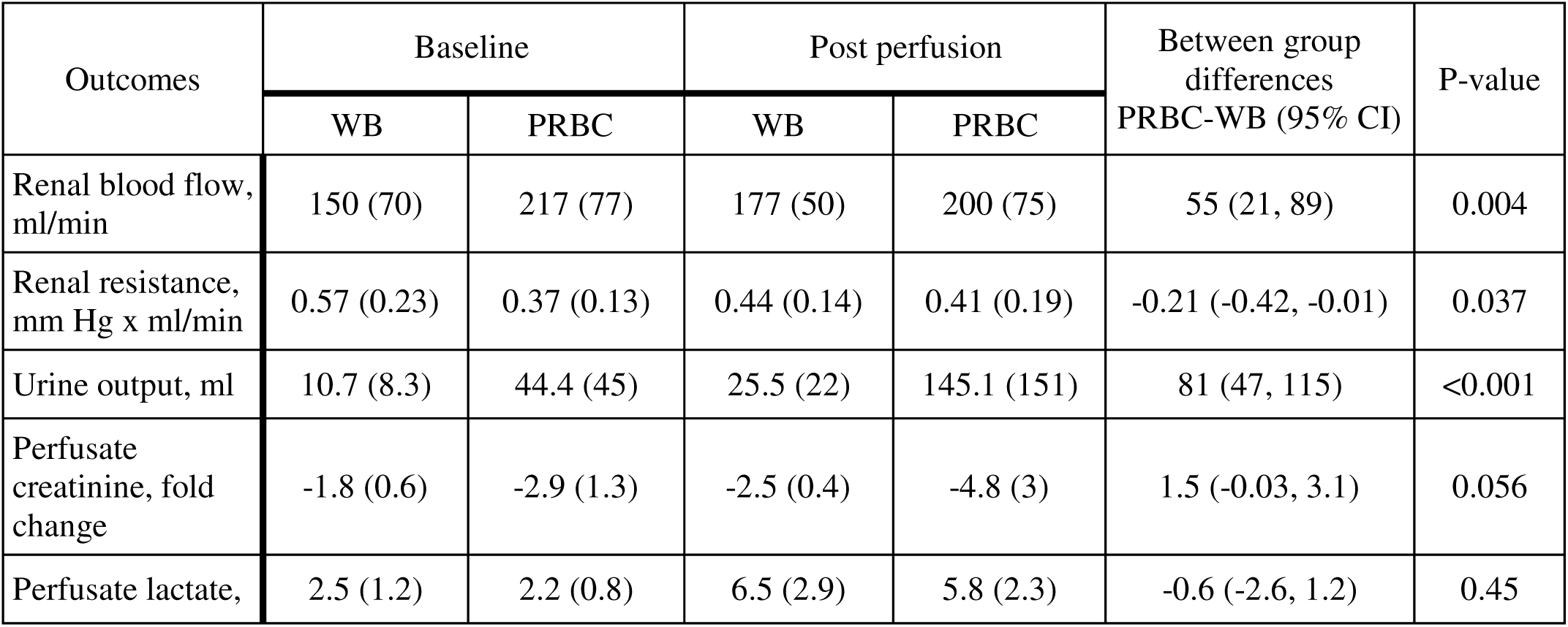
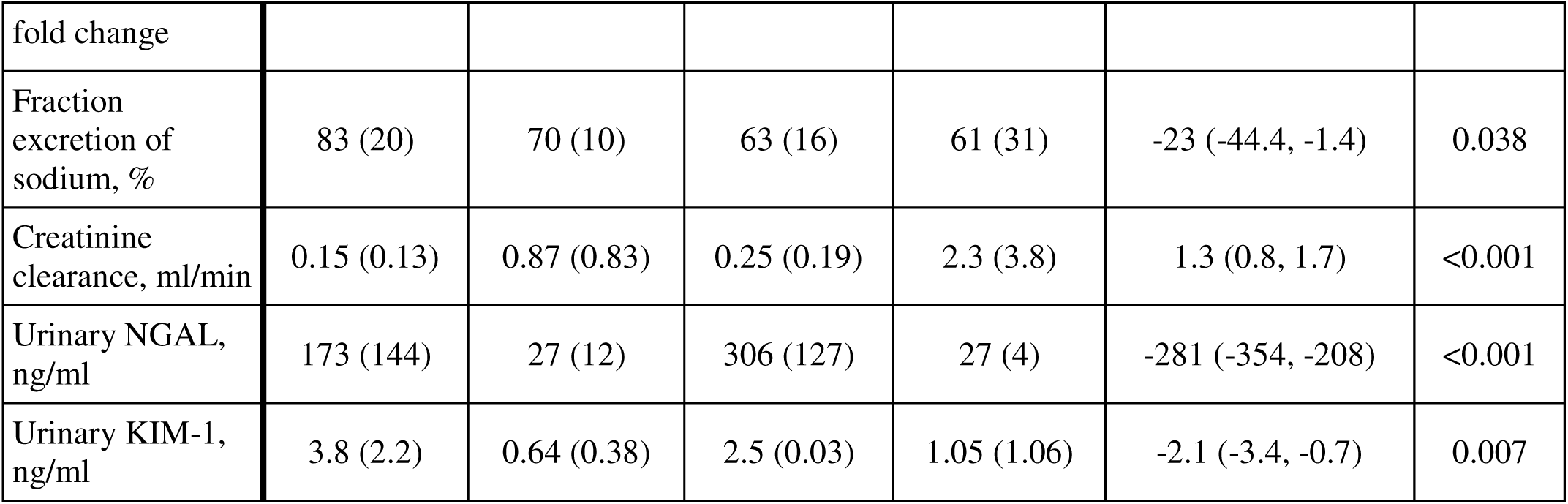
Summary of hemodynamics, functional, and injury biomarkers comparing WB and PRBC at baseline and post perfusion. Baseline values for urine output, perfusate creatinine and lactate, FeNA, creatinine clearance, urinary NGAL, and KIM-1 represent 30-minute after the initiation of perfusion. The values represent mean and standard deviation.

Overall, perfusate creatinine decreased by a mean fold change of 3.7 over three in both groups but declined faster in the PRBC group compared to the WB group (**Figure 1D, Table 2).** Perfusate lactic acid levels significantly for both WB and PRBC groups similarly (**Figure 1E, Table 2**). The PRBC perfused kidneys had a more stable and 23% lower FENa when compared to the WB perfused kidneys (**Figure 1F, Table 2**). Creatinine clearance was also higher in the PRBC kidneys over time than the WB kidneys (**Table 2**).

### Kidney biomarkers trajectories

Urinary NGAL concentrations were significantly lower in the PRBC perfused kidneys compared to the WB perfused kidneys (**Figure 1G, Table 2**). The WB perfused kidneys also exhibited a progressive increase trend in urinary NGAL levels over the perfusion period when compared to the PRBC perfused group. Urinary KIM-1 concentrations also showed statistically lower values and a more stable trend in the PRBC perfused kidneys than the WB perfused kidneys which had fluctuating values throughout the perfusion period (**Figure 1H, Table 2**).

### Histology

All biopsies taken at the end of the perfusion period showed widespread acute tubular injury with flattening of the tubular epithelia, dilation of tubular lumina, and irregular vacuolization of the tubular epithelial cytoplasm with no meaningful differences noted between the WB and PRBC perfused kidneys **(Table 1)**. Associated mild interstitial edema and scattered tubular cellular debris was noted as well (**Figure 2**).

**Figure 2.**
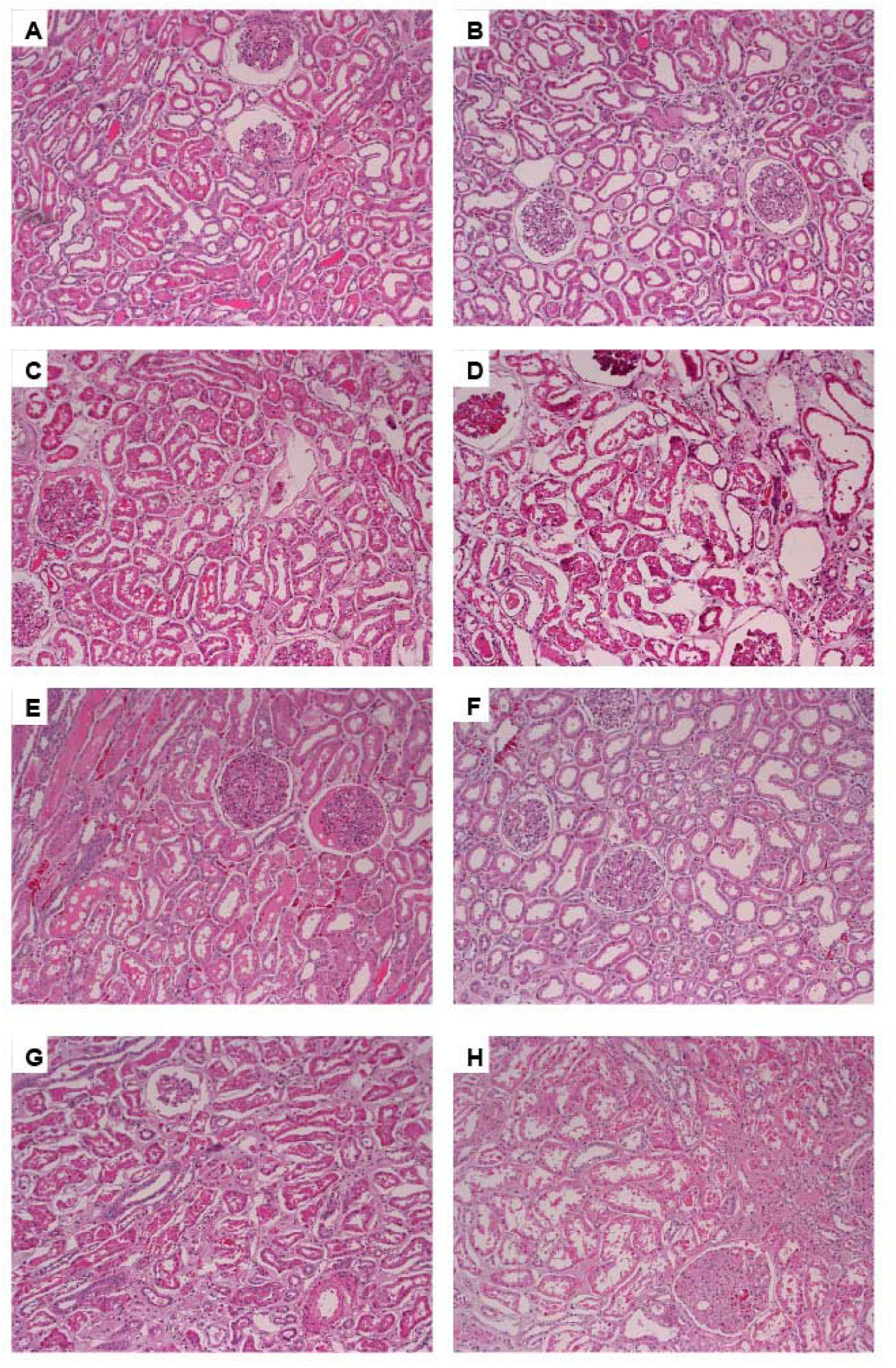
Histology after 3 hours of EVNP (100x). Renal cortical biopsies: H&E Stained, after 3 hours of EVNP with either whole blood (A, C, E, and G) or the paired kidney perfused with packed RBC based solution (B, D, F, H, respectively). All kidneys showed widespread acute tubular injury with flattening of the tubular epithelia, dilation of tubular lumina, and irregular vacuolization of the tubular epithelial cytoplasm. Scattered tubules contained tubular epithelial cells and cell debris. Associated mild interstitial edema is noted as well.

### Tissue Metabolic Changes

Before the initiation of perfusion tissue metabolic profile differences were minimal (1%) between groups. We observed marked tissue metabolic alterations in the WB group over time with a total of 43 (23%) metabolites compared to baseline (**Figure 3**). These temporal differences were predominantly characterized by heightened levels of (branched chain) amino acids (13/43 altered metabolites) and metabolites involved in mitochondrial energy metabolism. The metabolites with largest fold changes were glucose (9.8), ascorbic acid (9.2), and trehalose (7.6) (**Figure 3 and Supplemental Table 1**). Pathway analysis revealed amino acid and carbohydrate metabolism, and TCA cycle as the main altered metabolic pathways in the WB group during the 3-hour perfusion (**Supplemental Figure 1A**).

**Figure 3.**
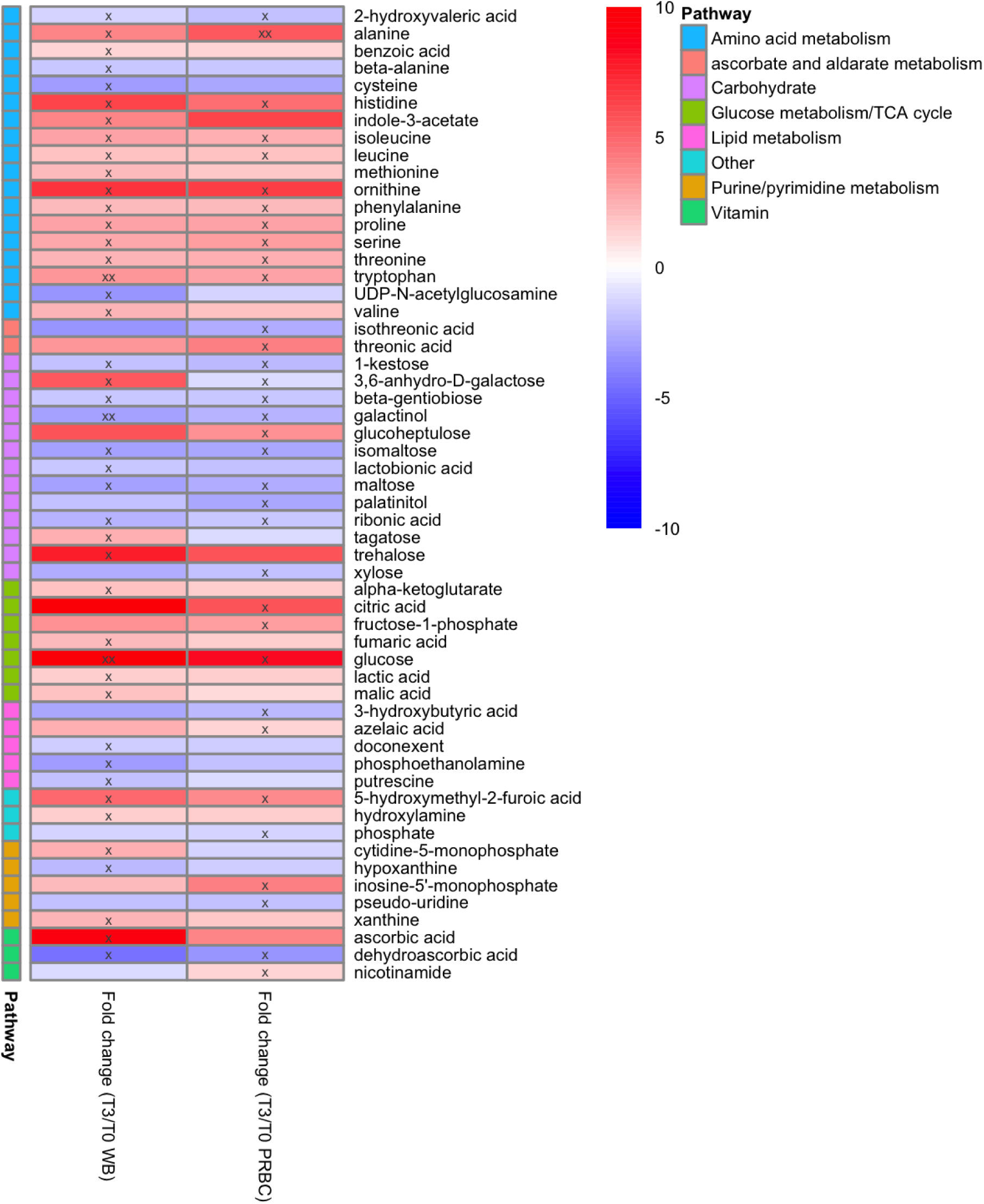
Tissue metabolic changes comparing pre vs post perfusion in the WB (n=4) and PRBC (n=4) groups. The fold changes are in reference to baseline. “x” represents a P <0.05 “xx” represents P <0.001.

In the PRBC group temporal tissue metabolic differences were noted for a total of 34 (18%) significant alteration comparing pre- and post- perfusion (**Figure 3**). Similar to the WB group, temporal differences were associated with elevated tissue levels of amino acids (10/34 altered metabolites) and carbohydrates. The metabolites with the largest fold changes over time were glucose (8.3), ornithine (6.4), and citric acid (5.6) (**Figure 3 and Supplemental Table 2**). Pathway analysis highlighted similar altered metabolic pathways to the WB group identifying (branched chain) amino acids and carbohydrate metabolism as the predominant altered pathways over time (**Supplemental Figure 1B**).

We found a remarkable overlap in the significantly altered tissue metabolites over time regardless of perfusion condition (WB or PRBC). Overall, changes in 20 metabolites were overlapping in both groups; however, the magnitude of differences were less pronounced in the PRBC compared to the WB group (**Figure 3**). The overlap predominantly highlighted aberrations involving amino acids and carbohydrate metabolism. Additionally, we identified differences in tissue metabolic changes from baseline to post perfusion comparing both groups. We found a total of 16 tissue metabolites to be significantly different comparing WB and PRBC groups over time (**Figure 4**). These metabolites included purine/pyrimidine metabolites, TCA cycle intermediates, and lipid peroxidation products. Metabolic pathways distinguishing the metabolic changes between the groups were amino acid (Arginine, Alanine, Aspartate, and Glutamate) metabolism, TCA cycle, and pyruvate metabolism (**Supplemental Figure 1C**).

**Figure 4.**
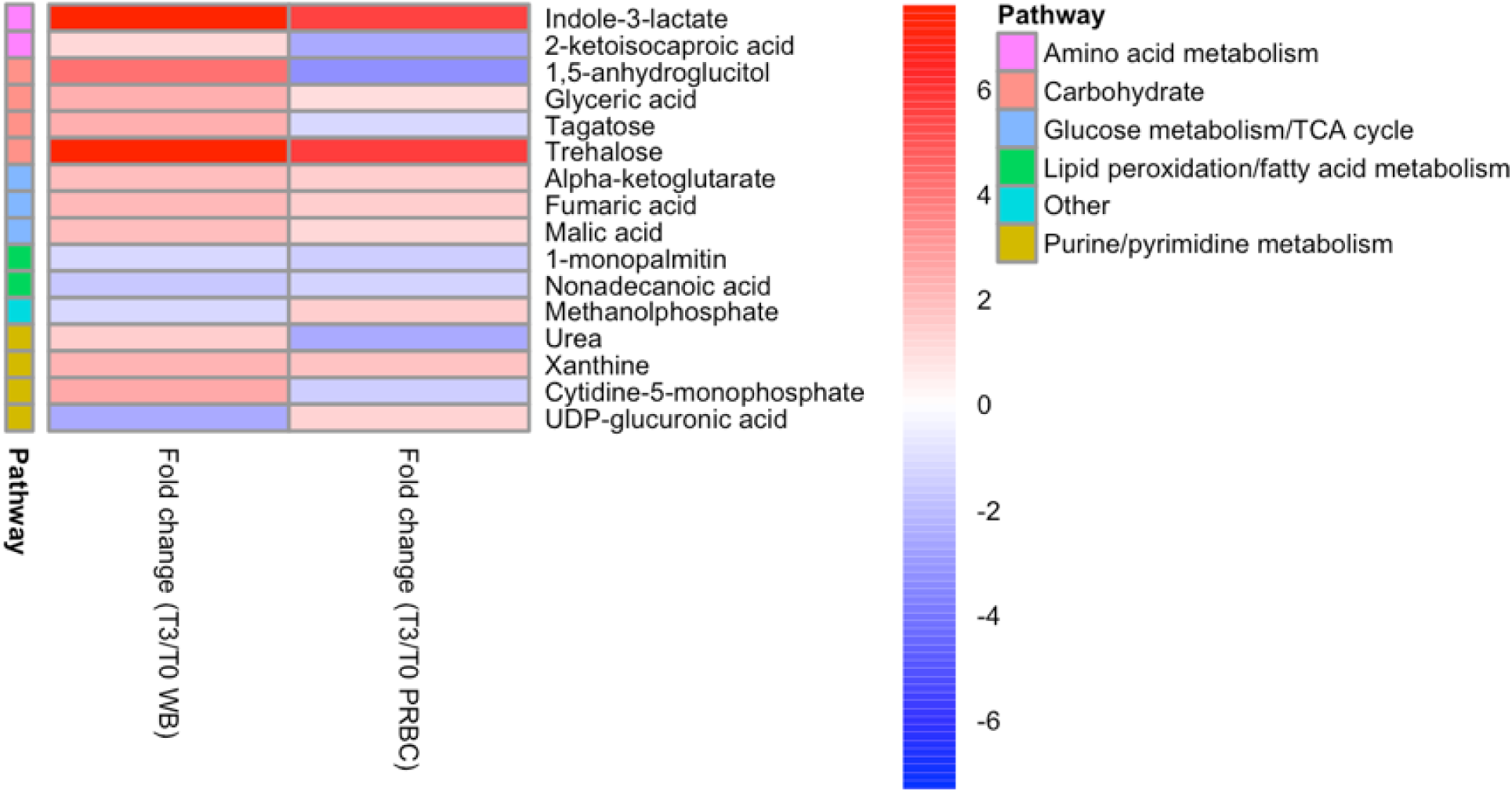
Tissue metabolic differences comparing WB (n=4) and PRBC (n=4) over time. The fold changes are in reference to baseline. Metabolites with interaction P <0.05 are shown.

### Tissue Lipid Profile Changes

In contrast to the minimal baseline difference in tissue metabolic profile between WB and PRBC groups, we noted meaningful baseline lipid profile differences including 34 (16%) significantly different lipid species distinguishing WB and PRBC groups (**Supplementary** Figure 2). All significantly different lipid species were lower in the WB group compared to PRBC. These lipid species were predominantly composed of glycerophospholipids followed by glycerolipids and sphingolipids (**Supplementary Table 1**).

Within group temporal lipid profile assessment revealed marked lipid species alterations in the WB group. Compared to baseline, a total of 48 (22%) tissue lipid species were significantly different post perfusion (**Figure 5**). The majority of the altered tissue lipids were elevated post perfusion compared to baseline levels. These lipids were mainly plasma membrane/structure components such as glycerolipids (TGs), glycerophospholipids, and steroids (**Figure 5 and Supplemental Table 3**).

**Figure 5.**
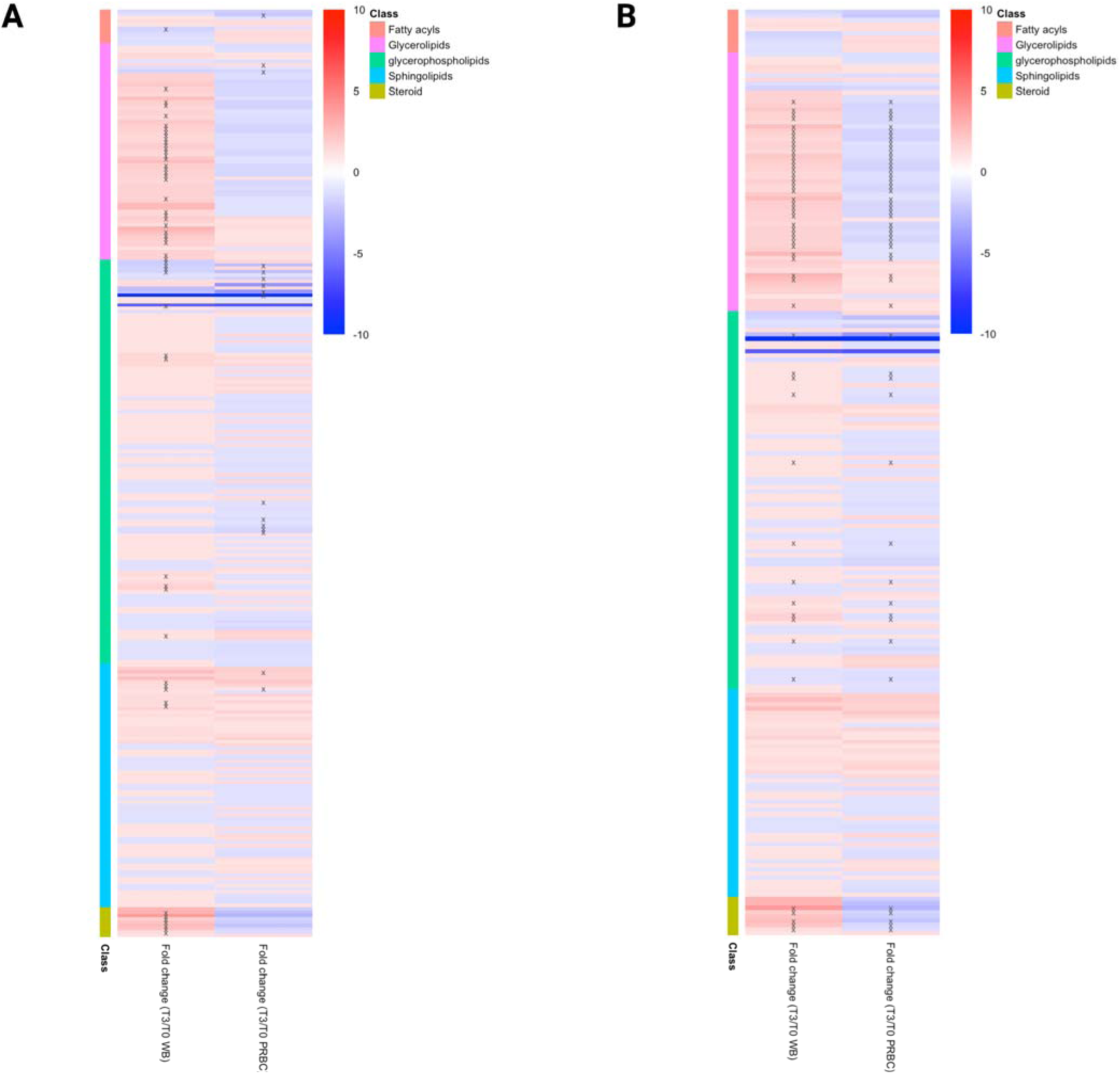
Tissue lipid profile differences pre and post perfusion within WB (n=4) and PRBC (n=4) groups and over time group comparisons. A) Tissue lipid profile within each group comparing pre and post perfusion. The fold changes are in reference to baseline. “x” represents a P <0.05 “xx” represents P <0.001. B) Tissue lipid profile differences comparing WB and PRBC over time. Lipids with interaction P <0.05 are shown.

The PRBC group demonstrated substantially fewer temporal differences in tissue lipid profile compared to the WB group. Only 13 (6%) of lipid species were significantly altered comparing pre- and post- perfusion (**Figure 5**). They predominantly consisted of glycerophospholipids. Interestingly, most of the altered lipids were lower post perfusion compared to baseline (**Figure 5 and Supplemental Table 4**).

A total of 53 (24%) lipid species were significantly different between the WB and PRBC groups over time. Unlike the PRBC group, the majority of these lipid species were elevated post-perfusion among the kidneys in the WB group (**Figure 5 and Supplemental Table 5**).

## Discussion

Despite the use of currently available modalities, many high-risk kidneys are discarded due to the inability to adequately assess the risk of poor outcomes prior to transplantation. Here, we used a three-hour EVNP system to assess hemodynamics, functional, and tissue metabolic changes among 8 paired high-risk deceased donor kidneys with relatively long CIT using PRBC or WB. Despite the similarity of WB and PRBC perfused kidneys on histology, kidneys perfused with PRBC had higher urine production and RBF and lower levels of urinary NGAL and KIM-1 suggesting improved viability. Metabolomics assessment during EVNP showed more drastic metabolic aberration in the WB group involving amino acid, carbohydrate, and mitochondrial energy metabolism. Tissue lipid profiling showed that WB was associated with accumulation of tissue membrane/structure components including TGs, ceramides, and cholesteryl esters post perfusion compared to the PRBC which demonstrated a minimal lipid profile change over time. Together, our findings reveal the superiority of PRBCs (leukocyte and platelet depletion) in facilitating and maintaining functional and metabolic recovery of marginal kidneys during an EVNP.

Our findings are consistent with prior studies showing that compared to whole blood leukocyte and platelet depleted blood based perfusate improved post-ischemic renal function after a period of warm ischemia^10,11^. Consistent with the studies from Hosgood^10,11^, we observed that the kidneys perfused with a PRBC perfusate exhibit superior kidney functional parameters compared the WB perfused kidneys evident by higher renal blood flow indicating superior vascular integrity (lower vascular injury and interstitial edema). We also observed evidence of higher glomerular filtration and tubular function with higher urine output, decreasing perfusate creatinine, and a lower fraction excretion of sodium of the PRBC group. Lastly, urinary inflammatory mediators and markers of proximal tubule injury were stable and consistently lower among the PRBC group compared to WB. Overall, our findings corroborate previous studies showing the role of leukocytes and platelets in exacerbating early kidney reperfusion injury resulting in impaired kidney function.

The relatively greater kidney function in the PRBC perfusate group compared to WB were observed in absence of any histologic differences between the two groups. In both groups, we observed comparable histological damage associated with reperfusion injury resulting in increased tubular dilation and vacuolation. This suggests that leukocyte and platelet depletion is not sufficient in mitigating tubular damage induced by early graft reperfusion as reported by others^17,18^. This is in line with previous studies comparing leukocyte depleted and WB perfusates of porcine kidneys^17,18^. Overall, our short-term post perfusion histological assessment did not agree with functional differences and therefore is not a sensitive early time point indicator for kidney function and recovery potential for high-risk kidneys.

Tissue metabolic profiling of the renal cortex has the advantage of providing biologic insight into the metabolic basis for kidney reperfusion injury, the influence of presence of leukocytes and platelets on metabolism during perfusion, and the association of metabolic health with kidney function. First, we found notable similarities in within group temporal changes among both groups mainly involving impairment in amino acids (and BCAAs) and glucose metabolism; however, these changes were more pronounced (higher fold changes of the same metabolites) among the WB group. Regardless of perfusion conditions, ischemia reperfusion injury is accompanied by metabolic aberrations associated and the presence of leukocytes may further influence glucose and amino acid metabolism. The proximal tubule actively reabsorbs nearly all of the filtered glucose and amino acid, incurring a high energetic demand from mitochondrial oxidative phosphorylation^19,20^. Second, the WB group was associated with diminished mitochondrial energy metabolism evidenced by impaired TCA cycle and pyruvate metabolism. Third, the differences in tissue metabolic profile were reflected in the hemodynamic and functional differences among the two groups. These findings are consistent with our previous study reporting that BCAA and glucose utilization are major determinants of kidney function and recovery in high-risk kidneys perfused using the same ENVP system^21^. Our findings also agree with another study that identified “amino acid transport” as one of the top altered metabolic pathways associated with delayed graft function from a cohort of 190 kidneys transplanted post-hypothermic perfusion^22^. Together, our data suggest that leukocyte depleted perfusion of high-risk kidneys mitigates reperfusion-associated metabolic derangements linked to mitochondrial dysfunction.

Leveraging practical and easy-to-measure tissue metabolic markers and simultaneous functional measures in the ex-vivo setting allows for a more comprehensive pre-transplant organ assessment approach to predict long term graft function and determine suitability for transplantation. In addition to amino acids and TCA cycle intermediates, we identified other tissue metabolites of interest that warrant further examination and validation to be used as such biomarkers. These metabolites include 1, 5 anhydroglucitol (1,5 AG), tagatose, and 2-ketoisocaproic acid (2-KIC) which were all elevated compared to their baseline level in the WB group. The clinical relevance of 1, 5 AG was recently underscored by a longitudinal analysis of 1612 adults with mean eGFR of 62 ml/min/1.73m^2^ from the Atherosclerosis Risk in Communities (ARIC) study showing that serum 1, 5 AG were associated with 40% decline in eGFR^23^. Additionally, tagatose has also been identified as a sensitive and specific metabolite for early diagnosis of acute ejection fraction after heart transplantation in preclinical models^24^. Lastly, 2-ketoisocaproic acid-the (abnormal) catabolic product of leucine catalyzed by the enzyme branched-chain alpha-keto acid dehydrogenase complex (BCKDC)^25^, can serve as a biomarker for impaired BCAA metabolism and mitochondrial health as is it localized in the inner mitochondrial membrane.

Tissue lipid profiling revealed marked differences between the WB and PRBC groups. We found more extensive lipid profile differences among the WB group. These lipids involved significant accumulation of plasma membrane/structure components such as glycerolipids (TGs), sphingolipids (ceramides), and steroids (cholesteryl esters). This elevation in cell membrane components suggests that increased tissue damage and necrosis resulted from ischemia damage in the presence of leukocytes and platelets. This is in line with our recent study showing early accumulation of plasma membrane lipids, including glycerolipids and sphingolipids, linked with poor kidney function and recovery during a 12-hour ENVP perfusion period^21^. Mechanistic studies support leukocyte depletion shown to reduce tubular apoptosis, caspase-3 activity, and IL-1β activation in porcine kidneys^14^. Together, our findings suggest that the presence of leukocytes further exacerbates ischemia injury contributing to poor kidney function possibly limiting recovery potential.

Our study had notable strengths and limitations. Using a well-established ENVP system, we performed a comprehensive organ assessment consisting of hemodynamic and functional measurements, histological analysis, and untargeted tissue metabolic/lipid profiling of high-risk kidneys. We evaluated 8 paired kidneys from four individuals allowing us to minimize between group variations in our assessments. Indeed, our study was not without limitations. Despite using 8 paired kidneys, our study cohort size was relatively small. The relatively long CIT of the kidneys (>50 hours) is at the extreme limit that many transplant centers would consider acceptable for transplantation. The perfusion duration was only limited to three hours, and it is possible that a longer perfusion period is necessary to adequately assess kidney viability and validate longer-term post-transplant outcomes. Lastly, lower viability combined with reduced clearance among the WB group might have led to passive diffusion of metabolites into the cells contributing to exaggerated tissue metabolite levels over time.

In conclusion, this study supports the potential role of leukocyte depleted EVNP system in evaluating and facilitating recovery of high-risk kidneys that are currently being discarded. Currently, there are no widely accepted EVNP criteria which definitely identify which high risk kidneys are safe to transplant. In addition to the previously established criteria for kidney transplantation such as macroscopic appearance, renal blood flow, and urine production^26,27^; early time-point tissue metabolic and lipid profile biomarkers (glycerolipids) should also be employed to enhance the predictability and success of the pre-transplant criteria. Additionally, longer duration studies with more comprehensive metabolic substrates are needed to fully assess the potential of the ENVP system in recovering marginal kidneys.

## Data Availability

All data produced in the present study are available upon reasonable request to the authors

## Abbreviations

CIT: cold ischemia time
EVNP: ex-vivo normothermic perfusion
FENa: fractional excretion of sodium
HMP: hypothermic machine perfusion
KDPI: kidney donor profile index
KIM-1: kidney injury molecule-1
NGAL: neutrophil gelatinase-associated lipocalin
PRBC: packed red blood cell
RBF: renal blood flow
RR: renal resistance
SCS: static cold storage
UO: urine output
WB: whole blood

## Acknowledgment

We would also like to thank the West Coast Metabolomics Center at University of California Davis for their contribution to this project.

## Author Contributions

The conceptualization was contributed by AA, HY, BR, and RVP. The methodology was contributed by AA, SF, and BR. The formal analysis was conducted by AA and SF. The investigation was performed by BR and RVP. Resources were contributed by BR and RVP. Data curation was performed by AA and SF. The original draft was written by AA, HY, IP, BR, and RVP. The review was written and edited by AA, HY, KJ, IP, JS, BR, and RVP. Visualization was contributed by AA. Supervision was carried out by BR and RVP. Project administration was contributed by BR and RVP. Funding acquisition was contributed by BR and RVP.

## Data Sharing Statement

A complete deidentified metadata supporting the findings in this study has been made available on Figshare (DOI: 10.6084/m9.figshare.26961916). Tissue metabolomics and lipidomics data can be found at (DOI: 10.6084/m9.figshare.26961934). Additional information will be made available to share upon request.

---

**Supplemental Table 1.** Tissue temporal metabolic differences among the WB group comparing pre and post perfusion (n=4).

| Metabolites | Fold change (T3/T0) | P-value | Pathway |
| --- | --- | --- | --- |
| Glucose | 9.8 | <0.01 | Sugar |
| Ascorbic acid | 9.2 | <0.01 | Vitamin |
| Trehalose | 7.6 | 0.03 | Sugar |
| Ornithine | 6.8 | <0.01 | Urea cycle |
| Histidine | 6.3 | <0.01 | Amino acid |
| 5-hydroxymethyl-2-furoic acid | 4.9 | <0.01 | nematicide |
| 2-hydroxyvaleric acid | 4.3 | 0.03 | Branched-chain amino acid metabolites (valine) |
| Indole-3-acetate | 3.9 | 0.04 | Tryptophan metabolism |
| Alanine | 3.9 | 0.04 | Amino acid |
| Tryptophan | 3.3 | <0.01 | Amino acid |
| Isoleucine | 2.9 | <0.01 | Amino acid |
| 3,6-anhydro-D-galactose | 2.9 | 0.01 | Sugar |
| Proline | 2.8 | 0.02 | Amino acid |
| Serine | 2.6 | <0.01 | Amino acid |
| Cytidine-5-monophosphate | 2.5 | 0.02 | Pyrimidine metabolism |
| Tagatose | 2.4 | 0.04 | Sugar |
| Threonine | 2.3 | <0.01 | Amino acid |
| Xanthine | 2.2 | 0.01 | purine metabolism |
| Valine | 2.2 | <0.01 | Amino acid |
| Phenylalanine | 2.1 | <0.01 | Amino acid |
| Methionine | 2.1 | 0.04 | Amino acid |
| Fumaric acid | 2.1 | <0.01 | TCA cycle |
| Malic acid | 1.9 | 0.01 | TCA cycle |
| Alpha-ketoglutarate | 1.9 | 0.04 | TCA cycle |
| Leucine | 1.8 | 0.01 | Amino acid |
| Lactic acid | 1.5 | 0.02 | Glycolysis/TCA |
| Hydroxylamine | 1.4 | 0.03 | Antioxidant |
| Benzoic acid | 1.3 | 0.02 | amino acid metabolism |
| Doconexent | 0.6 | <0.01 | alpha-Linolenic acid metabolism |
| Putrescine | 0.5 | 0.04 | Fatty acid metabolism |
| Lactobionic acid | 0.5 | 0.04 | Sugar |
| Beta-gentiobiose | 0.5 | 0.04 | Sugar |
| Beta-alanine | 0.5 | 0.02 | amino acid metabolism |
| 1-kestose | 0.5 | 0.04 | Sugar |
| Ribonic acid | 0.4 | <0.01 | Sugar acid |
| Hypoxanthine | 0.4 | 0.02 | purine metabolism |
| Phosphoethanolamine | 0.3 | 0.01 | Starch and sucrose metabolism |
| Maltose | 0.3 | 0.03 | Sugar |
| Isomaltose | 0.3 | <0.01 | Sugar |
| Galactinol | 0.3 | <0.01 | Galactose Metabolism |
| Cysteine | 0.3 | 0.01 | Amino acid |
| UDP-N-acetylglucosamine | 0.2 | 0.02 | amino acid metabolism |
| Dehydroascorbic acid | 0.2 | 0.02 | Vitamin |

**Supplemental Figure 1.**
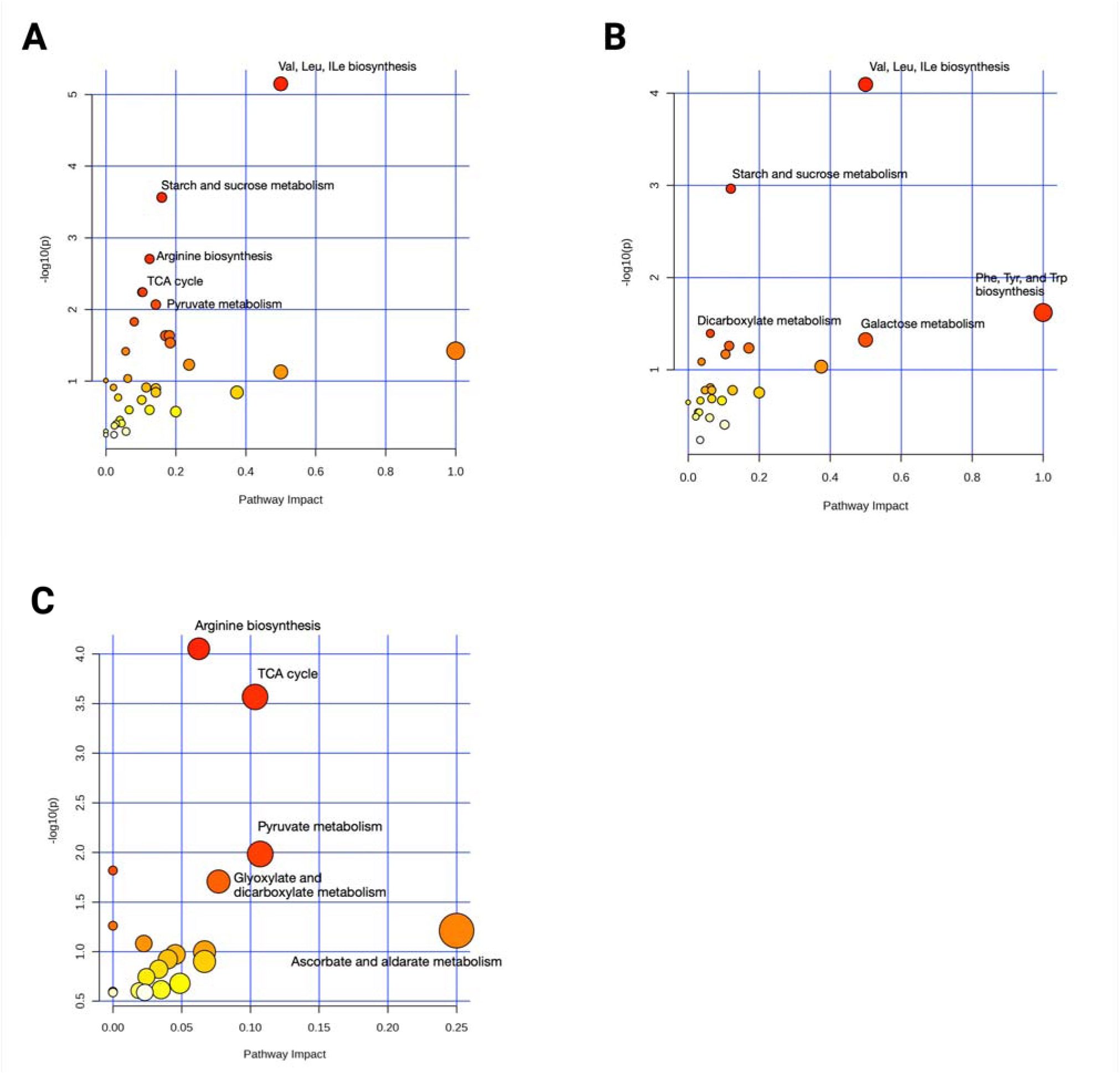
Metabolic pathway analysis of changes within WB/PRBC groups and temporal derangements comparing the two groups. A) Pre vs post WB, B) pre vs post PRBC, and C) WB vs PRBC over time. The size and color of the nodes represent pathway impact value and *P* value, respectively.

**Supplemental Table 2.** Tissue temporal metabolic differences among the PRBC group comparing pre and post perfusion (n=4).

| Metabolites | Fold change (T3/T0) PRBC | P-value | Pathway |
| --- | --- | --- | --- |
| Glucose | 8.3 | <0.01 | Sugar |
| Ornithine | 6.4 | <0.01 | Urea cycle |
| 2-hydroxyvaleric acid | 5.6 | 0.03 | Branched-chain amino acid metabolites (valine) |
| Citric acid | 5.6 | <0.01 | TCA cycle |
| Alanine | 5.4 | <0.01 | Amino acid |
| Histidine | 4.7 | 0.02 | Amino acid |
| Inosine-5'-monophosphate | 4.0 | 0.02 | Purine metabolism |
| Threonic acid | 4.0 | 0.04 | ascorbate and aldarate metabolism |
| 5-hydroxymethyl-2-furoic acid | 3.7 | <0.01 | nematicide |
| 3,6-anhydro-D-galactose | 3.6 | 0.01 | Galactose metabolism |
| Glucoseheptulose | 3.4 | <0.01 | Sugar |
| Fructose-1-phosphate | 3.1 | 0.02 | Glycolysis |
| Serine | 3.0 | <0.01 | Amino acid |
| Proline | 2.9 | <0.01 | Amino acid |
| Tryptophan | 2.9 | 0.04 | Amino acid |
| 3-hydroxybutyric acid | 2.5 | <0.01 | Fatty acid metabolism |
| Isoleucine | 2.5 | 0.02 | Amino acid |
| Threonine | 2.4 | <0.01 | Amino acid |
| Phenylalanine | 2.0 | 0.04 | Amino acid |
| Leucine | 1.9 | 0.03 | Amino acid |
| Azelaic acid | 1.3 | 0.01 | Fatty acid metabolism |
| Nicotinamide | 1.3 | 0.03 | Vitamin |
| Phosphate | 0.8 | <0.01 | Electrolyte |
| Beta-gentiobiose | 0.5 | 0.04 | Sugar |
| Pseudo-uridine | 0.5 | 0.04 | Pyrimidine metabolism |
| Ribonic acid | 0.5 | <0.01 | Sugar acid |
| Xylose | 0.5 | <0.01 | Sugar |
| 1-kestose | 0.4 | 0.03 | Sugar |
| Galactinol | 0.4 | 0.03 | Galactose Metabolism |
| Dehydroascorbic acid | 0.3 | 0.02 | Vitamin |
| Isomaltose | 0.3 | 0.02 | Sugar |
| Isothreonic acid | 0.3 | 0.04 | ascorbate and aldarate metabolism |
| Maltose | 0.3 | 0.04 | Sugar |
| Palatinitol | 0.3 | 0.01 | Sugar |

**Supplemental Figure 2.**
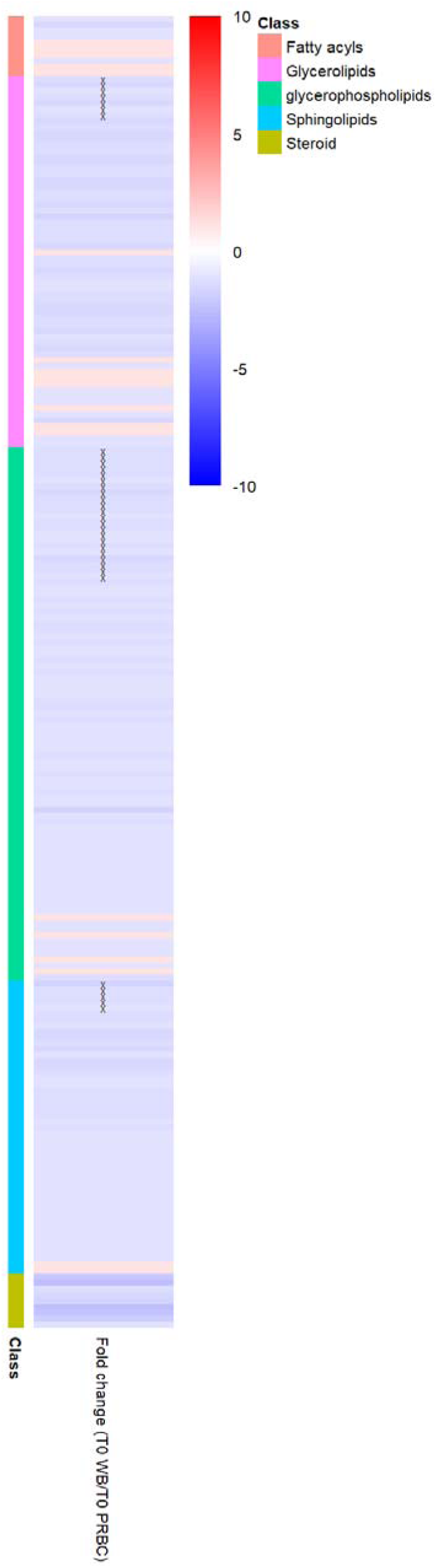
Tissue lipid baseline differences comparing WB and PRBC. “x” represents a P <0.05 “xx” represents P <0.001.

**Supplemental Table 3.** Tissue temporal lipid profile differences among the WB group comparing pre and post perfusion (n=4).

**profile differences among the WB group comparing pre and post perfusion (n=4).**
| Lipids | Fold change (T3/T0 WB) | P-value | Lipid class |
| --- | --- | --- | --- |
| CE (18:2) | 6.3 | 0.03 | Steroids |
| CE (20:5) | 3.4 | <0.01 | Steroids |
| TG (56:5) | 3.4 | <0.01 | Glycerolipids |
| CE (18:1) | 3.3 | 0.04 | Steroids |
| TG (48:0) | 3.3 | 0.01 | Glycerolipids |
| TG (54:6) | 3.3 | 0.01 | Glycerolipids |
| TG (52:1) | 3.2 | 0.04 | Glycerolipids |
| CE (20:3) | 3.1 | 0.04 | Steroids |
| CE (20:4) | 3.0 | 0.04 | Steroids |
| TG (50:1) | 2.9 | 0.01 | Glycerolipids |
| TG (48:1) | 2.7 | 0.01 | Glycerolipids |
| TG (50:2) | 2.6 | 0.01 | Glycerolipids |
| TG (52:2) | 2.6 | 0.03 | Glycerolipids |
| TG (56:6) | 2.6 | <0.01 | Glycerolipids |
| TG (46:0) | 2.5 | 0.04 | Glycerolipids |
| TG (50:3) | 2.5 | 0.04 | Glycerolipids |
| TG (52:3) | 2.4 | 0.02 | Glycerolipids |
| TG (50:4) | 2.3 | 0.03 | Glycerolipids |
| TG (51:1) | 2.3 | 0.03 | Glycerolipids |
| TG (54:4) | 2.2 | 0.03 | Glycerolipids |
| CE (18:3) | 2.1 | 0.04 | Steroids |
| TG (51:3) | 2.1 | 0.03 | Glycerolipids |
| TG (52:4) | 2.1 | 0.02 | Glycerolipids |
| TG (52:5) | 2.1 | 0.02 | Glycerolipids |
| TG (56:7) | 2.1 | <0.01 | Glycerolipids |
| TG (56:8) | 2.1 | 0.03 | Glycerolipids |
| TG (51:2) | 2.0 | 0.03 | Glycerolipids |
| TG (49:0) | 1.9 | 0.04 | Glycerolipids |
| TG (58:9) | 1.9 | 0.03 | Glycerolipids |
| TG (50:5) | 1.8 | 0.01 | Glycerolipids |
| TG (50:5) | 1.8 | <0.01 | Glycerolipids |
| TG (56:2) | 1.8 | 0.04 | Glycerolipids |
| TG (58:4) | 1.8 | 0.01 | Glycerolipids |
| Ceramide (d38:1) | 1.7 | 0.01 | Sphingolipids |
| PC (p-42:4) or PC (o-42:5) | 1.7 | 0.03 | Glycerophospholipids |
| PC (p-44:4) or PC (o-44:5) | 1.7 | <0.01 | Glycerophospholipids |
| CE (22:6) | 1.6 | 0.04 | Steroids |
| TG (54:7) | 1.5 | 0.03 | Glycerolipids |
| Ceramide (d40:1) | 1.4 | 0.01 | Sphingolipids |
| Ceramide (d40:1) | 1.4 | <0.01 | Sphingolipids |
| Ceramide (d42:2) | 1.4 | 0.02 | Sphingolipids |
| PC (p-40:3) or PC (o-40:4) | 1.3 | 0.01 | Glycerophospholipids |
| PE (p-36:1) or PE (o-36:2) | 1.2 | 0.04 | Glycerophospholipids |
| PC (34:0) | 1.1 | 0.03 | Glycerophospholipids |
| docosahexaenoic acid | 0.6 | 0.01 | Fatty acyls |
| LPC (16:0) | 0.6 | 0.04 | Glycerophospholipids |
| LPC (16:0) | 0.5 | 0.02 | Glycerophospholipids |
| LPC (20:4) | 0.2 | 0.03 | Glycerophospholipids |

**Supplemental Table 4.** Minimal temporal differences in tissue lipid profile among PRBC perfused kidneys (n=4).

|  | Fold Change<br>T3/T0 PRBC | P-value | Lipid class |
| --- | --- | --- | --- |
| Ceramide (d34:0) | 1.9 | 0.02 | Sphingolipids |
| DG (38:3) | 1.4 | 0.04 | Glycerolipids |
| Ceramide (d40:1) | 1.1 | 0.04 | Sphingolipids |
| PC (38:5) | 0.8 | 0.01 | Glycerophospholipids |
| PC (40:6) | 0.8 | 0.03 | Glycerophospholipids |
| DG (38:6) | 0.6 | 0.04 | Glycerolipids |
| PC (40:7) | 0.6 | <0.01 | Glycerophospholipids |
| PC (40:8) | 0.6 | 0.01 | Glycerophospholipids |
| LPC (18:0) | 0.5 | 0.01 | Glycerophospholipids |
| Acylcarnitine C18:0 | 0.4 | <0.01 | Fatty acyls |
| LPC (16:0) | 0.4 | 0.03 | Glycerophospholipids |
| LPC (18:1) | 0.3 | 0.03 | Glycerophospholipids |
| LPC (18:2) | 0.1 | 0.03 | Glycerophospholipids |

**Supplemental Table 5.** Differences in tissue lipid profile comparing WB (n=4) and PRBC (n=4) over time.

|  | Fold change<br>(T3/T0)WB | Fold change<br>(T3/T0)PRBC | P-value for<br>interaction | Lipid Class |
| --- | --- | --- | --- | --- |
| CE (18:2) | 3.987 | 0.341 | 0.044 | Steroids |
| TG (56:5) | 2.985 | 1.206 | 0.048 | Glycerolipids |
| TG (54:6) | 2.679 | 0.889 | 0.017 | Glycerolipids |
| CE (20:4) | 2.550 | 0.449 | 0.048 | Steroids |
| TG (56:6) | 2.437 | 1.148 | 0.040 | Glycerolipids |
| TG (48:0) | 2.423 | 0.836 | 0.012 | Glycerolipids |
| TG (52:1) | 2.422 | 0.967 | 0.035 | Glycerolipids |
| CE (20:5) | 2.234 | 0.670 | 0.019 | Steroids |
| TG (46:0) | 2.158 | 0.697 | 0.029 | Glycerolipids |
| TG (50:1) | 2.107 | 0.741 | 0.009 | Glycerolipids |
| CE (18:3) | 1.973 | 0.611 | 0.038 | Steroids |
| TG (50:2) | 1.921 | 0.670 | 0.013 | Glycerolipids |
| TG (48:1) | 1.896 | 0.588 | 0.019 | Glycerolipids |
| TG (51:1) | 1.867 | 0.774 | 0.019 | Glycerolipids |
| TG (52:2) | 1.860 | 0.691 | 0.006 | Glycerolipids |
| TG (52:3) | 1.810 | 0.651 | 0.007 | Glycerolipids |
| TG (49:0) | 1.796 | 0.976 | 0.010 | Glycerolipids |
| TG (44:1) | 1.777 | 0.791 | 0.036 | Glycerolipids |
| TG (54:2) | 1.746 | 0.815 | 0.039 | Glycerolipids |
| TG (46:1) | 1.736 | 0.646 | 0.027 | Glycerolipids |
| PC (p-42:4) or PC (o-42:5) | 1.703 | 0.997 | 0.020 | Glycerophospholipids |
| TG (46:2) | 1.701 | 0.683 | 0.039 | Glycerolipids |
| TG (52:4) | 1.700 | 0.676 | 0.010 | Glycerolipids |
| TG (53:4) | 1.669 | 0.665 | 0.009 | Glycerolipids |
| TG (50:3) | 1.667 | 0.583 | 0.022 | Glycerolipids |
| PC (p-44:4) or PC (o-44:5) | 1.659 | 0.923 | 0.018 | Glycerophospholipids |
| TG (54:5) | 1.655 | 0.841 | 0.008 | Glycerolipids |
| TG (54:3) | 1.652 | 0.697 | 0.015 | Glycerolipids |
| TG (49:1) | 1.630 | 0.770 | 0.021 | Glycerolipids |
| TG (54:4) | 1.628 | 0.675 | 0.018 | Glycerolipids |
| TG (48:2) | 1.614 | 0.582 | 0.035 | Glycerolipids |
| TG (52:5) | 1.607 | 0.673 | 0.008 | Glycerolipids |
| TG (50:4) | 1.605 | 0.619 | 0.018 | Glycerolipids |
| TG (58:4) | 1.604 | 1.160 | 0.040 | Glycerolipids |
| TG (51:2) | 1.594 | 0.711 | 0.037 | Glycerolipids |
| TG (51:3) | 1.593 | 0.627 | 0.030 | Glycerolipids |
| TG (53:3) | 1.577 | 0.638 | 0.044 | Glycerolipids |
| TG (50:5) | 1.533 | 0.818 | 0.004 | Glycerolipids |
| TG (48:3) | 1.526 | 0.593 | 0.036 | Glycerolipids |
| TG (50:5) | 1.519 | 0.780 | 0.015 | Glycerolipids |
| TG (49:2) | 1.463 | 0.746 | 0.036 | Glycerolipids |
| TG (54:7) | 1.462 | 0.861 | 0.019 | Glycerolipids |
| CE (22:6) | 1.391 | 0.715 | 0.032 | Steroids |
| PC (p-40:3) or PC (o-40:4) | 1.303 | 0.899 | 0.018 | Glycerophospholipids |
| PC (33:0) | 1.169 | 0.903 | 0.007 | Glycerophospholipids |
| PC (p-34:0) or PC (o-34:1) | 1.108 | 0.918 | 0.039 | Glycerophospholipids |
| PC (36:1) | 1.102 | 0.863 | 0.024 | Glycerophospholipids |
| PC (32:0) | 1.060 | 0.921 | 0.030 | Glycerophospholipids |
| PE (36:1) | 1.030 | 0.864 | 0.010 | Glycerophospholipids |
| PC (40:4) | 1.029 | 0.755 | 0.005 | Glycerophospholipids |
| PC (31:0) | 1.021 | 0.861 | 0.043 | Glycerophospholipids |
| PE (p-38:4) or PE (o-38:5) | 0.957 | 0.829 | 0.045 | Glycerophospholipids |
| LPC (18:1) | 0.398 | 0.244 | 0.003 | Glycerophospholipids |

